# Role of Chloroquine and Hydroxychloroquine in the Treatment of COVID-19 Infection- A Systematic Literature Review

**DOI:** 10.1101/2020.03.24.20042366

**Authors:** Krishan Mohan Kapoor, Aanandita Kapoor

## Abstract

**Background:** Coronavirus pandemic is currently a global public health emergency. At present, no pharmacological treatment is known to treat this condition, and there is a need to review the available treatments.

**Objective:** While there have been studies to describe the role of chloroquine and hydroxychloroquine in various viral conditions, there is limited information about the use of them in COVID-19. This systematic review aims to summarize the available evidence regarding the role of chloroquine in treating coronavirus infection.

**Methods:** The preferred reporting items for systematic reviews and meta-analyses (PRISMA) guidelines were used for this review. A literature search was performed using PUBMED & Google Scholar to find articles about the role of CQ in COVID-19 patients.

**Results:** We included 19 publications (Five published articles, three letters/correspondence, one commentary, five pre-proofs of accepted articles, one abstract of yet to be published article, and four were pre-prints (not yet peer-reviewed) articles) in this systematic review. All the articles mentioned about the role of chloroquine and /or hydroxychloroquine in limiting the infection with SARS-CoV-2 (the virus causing COVID-19).

**Conclusions:** There is theoretical, experimental, preclinical and clinical evidence of the effectiveness of chloroquine in patients affected with COVID-19. There is adequate evidence of drug safety from the long-time clinical use of chloroquine and hydroxychloroquine in other indications. More data from ongoing and future trials will add more insight into the role of chloroquine and hydroxychloroquine in COVID-19 infection.

## Introduction

An outbreak of Severe Acute Respiratory Syndrome Coronavirus 2 (SARS-CoV-2 or COVID-19) was reported in Wuhan, China, in December 2019, which spread to the rest of China and many other countries across the globe. Globally the number of people diagnosed with COVID-19 infection is increasing exponentially, and it has been declared a pandemic by WHO. At present, no pharmacological agent has been approved by regulatory agencies for the treatment of SARS-CoV-2 infection, and a review of drugs found to be effective against COVID-19 is the need of the hour. Chloroquine (CQ) has been used extensively as an antimalarial agent with immunomodulatory effects^1 2 3^. A derivative of CQ, Hydroxychloroquine sulfate (HCQ) was synthesized first in 1946 by adding a hydroxyl group to CQ and is much less toxic than CQ in animal studies^4^. CQ has been used in SARS Coronavirus infection due to its antiviral properties^5 6^. Recently CQ has also been found to have Anti-COVID-19 activity in vitro^7^. For these reasons, CQ and HCQ could be the potential drug for treating COVID-19 infection. To date, there is no clinical evidence to support the use of CQ or HCQ for treating SARS-CoV-2 infection though many clinical trials with these drugs are already underway^8^. This study aims to systematically review the available literature for the use of CQ or HCQ in treating COVID-19 infection.

## Materials and Methods

A systematic review of the published literature was done to find the role of CQ in COVID 19 infection. The Preferred Reporting Items for Systematic Reviews and Meta-analyses, PRISMA, guidelines were used for the review. The publications selected in this study included the abstracts, original articles, pre-proofs of the accepted article, pre-prints, case reports or case series, letter to editor and experts’ opinions, published since inception till 23rd March, 2020, which investigated or discussed the role of CQ in COVID 19 infection. The articles excluded from this study were duplicate papers, posters, and papers without the mention of the role of CQ in COVID-19 infection.

### Literature search and Data Sources

PubMed and Google Scholar were used for searching the articles, reporting on the topics of the role of CQ in COVID 19 infection. The search was conducted using the following keywords: “Chloroquine,” “Hydroxychloroquine,” “COVID-19,” and “SARS-CoV-2” with a publication time range up to 23rd March, 2020. The abstract & purpose of the articles found during the literature search were reviewed. Articles describing the role of CQ or HCQ in COVID-19 infection were selected for full-text review.

### Screening

Titles, as well as the abstracts of items in the search results, were screened. The full-text of the selected articles was downloaded. Relevant information about the use of CQ and HCQ from all the selected articles was extracted. The data from selected articles were studied independently by two investigators (KMK and AK), and consensus was achieved with mutual discussion.

### Data Extraction and Analysis

The relevant data were extracted from selected articles by two authors (KMK and AK). The data points were tabulated after reading the full text of each selected article. As the number of the selected articles was small, the findings were presented in the form of a table & narrative summaries.

## Results

The initial search for the role of chloroquine in Covid-19 yielded 282 entries, published till 23^rd^ March, 2020. After using the exclusion criteria, eighteen items were selected. The list included five published articles^9 10 11 12 13^, three letters/correspondence^14 15 7^, one commentary^16^, four pre-proofs of accepted articles^17 18 19 20 21^, one abstract of yet to be published article^22^ and four w pre-prints (not yet peer-reviewed) articles^23 24 25 8^. One publication in Chinese^13^ was selected after a review of the English abstract, and it was translated in the English language using Google Translator service.

## Discussion

Chloroquine (CQ) is a well-known 4-aminoquinoline that has been in clinical use since 1944. Hydroxychloroquine sulfate (HCQ) was synthesized in 1946 with the addition of a hydroxyl group to CQ. CQ has been used extensively in treating malaria and also for the treatment of autoimmune disorders such as rheumatoid arthritis, systemic lupus erythematosus, etc. because of its immunomodulatory role^26 27^.

CQ exerts its antiviral effects through various mechanisms in the cell. CQ is weakly alkaline in nature, and it can change the pH of endosomes. Thus it can exert a significant inhibitory effect on viral infections, which invade cells via the endosome pathway like Zika virus^28 29^ and Borna disease virus^30^.

Many other viruses like Ebola (EBOV) depend on the naturally low pH of acidic endosomes for activating and triggering the fusion by their envelope glycoproteins. Thus a ‘fusion inhibitor’ could target the host cell machinery by preventing the acidification of the endosome to inhibit virus entry. The cell entry by EBOV, FLU-H5, and MARV can be inhibited due to the increase in the endosome pH by CQ due to its ability to prevent the acidification of intracellular organelles^31^.

The data on the kinetics of chloroquine uptake indicated the drug attained near-maximum intracellular concentrations in 1 hr and a drug level of 60 μg/ml produced a decrement in the cellular biosynthesis of protein, RNA, or DNA. Herpes simplex replication was found to be sensitive to this drug as the level of progeny virus production was depressed by 95% in 24 hr.^32^

CQ also affects viral replication by inhibiting viral gene expression. CQ has an ability to change the pattern of glycosylation in HIV-1 gp120 envelope during In vitro and in vivo tests. CQ also inhibits the replication of the HIV virus in CD4 + T cells^33^. Both CQ and HCQ are weakly alkaline bases and affect the acid vesicles causing the dysfunction of enzymes required for post-translational protein modifications. These drugs increase the pH of acidic vesicles, cause disruption of several enzymes like acid hydrolases & lead to inhibition of the post-transcriptional modification of newly synthesized proteins. The mechanisms thought to explain the anti-HIV-1 effect of CQ and HCQ include: alteration in gp120 production, restriction of intracellular iron (a cofactor) necessary for HIV-1 replication, an effect on Tat-mediated transactivation of the HIV-1 LTR, and the effect on the HIV-1 integrase.^34^

During animal testing, CQ was also found to effectively inhibit autophagy in the lungs of avian influenza H5N1 mice and reduce alveolar epithelial damage. CQ could efficiently ameliorate the acute lung injury and significantly improve the survival rate in mice infected with live avian influenza A H5N1 virus.^35^

Coronaviruses are enveloped viruses having non-segmented, single-stranded & positive-sense RNA genomes. Apart from causing infections in vertebrates in the meat industry like pigs and chickens, until recently, six coronaviruses are known to infect human hosts & cause respiratory diseases. COVID-19 is the latest addition to this family of viruses. Among these, severe acute respiratory syndrome coronavirus (SARS-CoV), Middle East respiratory syndrome coronavirus (MERS-CoV), and severe acute respiratory syndrome coronavirus 2 (SARS-CoV-2) are highly pathogenic coronaviruses which have resulted in regional and global outbreaks.^36^ Currently, there are no clinically specific pharmacologic agents for these seven coronaviruses.

CQ has been found to cause inhibition of the replication of SARS-CoV in Vero E6 cells. The IC_50_ of chloroquine inhibition of SARS-CoV replication in Vero E6 cells, 8.8 μM, is 1000-fold below the human plasma concentrations of CQ, reached following acute malaria treatment with chloroquine at a dose of 25 mg/kg over three days. The dose of chloroquine used for the treatment of rheumatoid arthritis (3.6 mg/kg) was found to produce plasma CQ concentrations of 1–3 μM, which is in the same concentration range as the IC_50_ for inhibition of SARS-CoV.^37 38^

CQ has been found to be effective in inhibiting the infection and spread of SARS CoV in cell culture. CQ inhibits the virus replication by reducing the terminal glycosylation of angiotensin-converting enzyme 2 (ACE2) receptors on the Vero E6 cells’ surface and interfering with the binding of SARS-CoV to the ACE2 receptors. CQ had a significant inhibitory antiviral effect when the susceptible cells were treated either before or after infection, suggesting a possible prophylactic and therapeutic use.^5^ CQ interferes with COVID-19’s attempt to acidify the lysosomes & probably inhibits cathepsins, which needs a low pH for optimal cleavage of COVID-19 spike protein, a requirement for the formation of the autophagosome.^18^

The Wuhan CoV (COVID-19) S-protein shares an almost identical 3-D structure in its RBD domain with SARS-CoV S-protein. Although relatively weaker than SARS-CoV, the Wuhan CoV S-protein has a strong binding affinity to human ACE2 receptors. Hence, the Wuhan CoV poses a significant risk for human transmission due to the S-protein–ACE2 binding pathway like SARS-CoV. ^20 39^

CQ has been found to function at both entry as well as post-entry stages of the COVID-19 infection in Vero E6 cells. The immune-modulating activity of CQ may also synergistically enhance its antiviral effect in vivo. After oral intake, CQ gets widely distributed in the whole body, including lung. The EC90 value of chloroquine against the COVID-19 in Vero E6 cells was 6.90μM, which is achievable clinically, as shown in the plasma of rheumatoid arthritis patients receiving 500 mg daily dose^7^. The low cost and good safety profile makes CQ a potentially clinically applicable drug against the COVID-19 infection.^7^

The endemic malaria presence seems to protect certain populations from COVID-19 infection, especially in the least developed countries. The mechanism of action of some antimalarial drugs with antiviral property suggests their potential role in the chemoprophylaxis of the COVID-19 epidemic.^22^ In a study, CQ has been found out to be the potential inhibitor of viral PLpro. The effect of chloroquine during post-entry stages could be manifested via its inhibition of the crucial viral protein, PLpro.^23^

The suggested regimen by Dutch CDC in adults includes an initial dose of 600 mg of chloroquine base followed by 300 mg after 12 h on the first day, followed by 300 mg twice daily on days second to fifth. This document also emphasizes the need for stopping the treatment at day 5 to reduce the risk of side effects, as CQ has a long half-life of 30 hours. There is also a need to differentiate between treatment based on chloroquine phosphate and chloroquine base since 500 mg of chloroquine phosphate is equivalent to 300 mg of chloroquine base. Another set of guidelines by the Italian Society of Infectious and Tropical disease (Lombardy section) suggests the use of CQ 500 mg twice daily or HCQ 200 mg once daily for ten days with treatment duration varying between 5 to 20 days based on clinical severity. The suggested target group of patients ranged from patients with mild respiratory symptoms having comorbidities to patients with severe respiratory failure.^11^

CQ has been recommended for 18–65 years of COVID-19 infected adults at a dose of 500mg twice daily for seven days. For BW ≤ 50 kg patients, the CQ dose should be decreased to 500 once daily during day third to seventh of the treatment cycle (Table 1). There has been no dose recommendation of CQ in COVID-19 infected children so far; hence extreme caution should be followed while prescribing it for children. Acute poisoning of CQ is usually fatal with a dose of 50 mg/kg, and CQ concentration > 25 μmol/L is a fatal predictor.^12^ In Nigerian children with acute uncomplicated malaria, chloroquine absorption appeared reliable as peak chloroquine concentrations were achieved within 0.5 hours, and its concentration decline slowly, with an elimination half-life of 20 to 60 days. CQ can be detected in urine months after a single dose.^40^

**Table 1:**
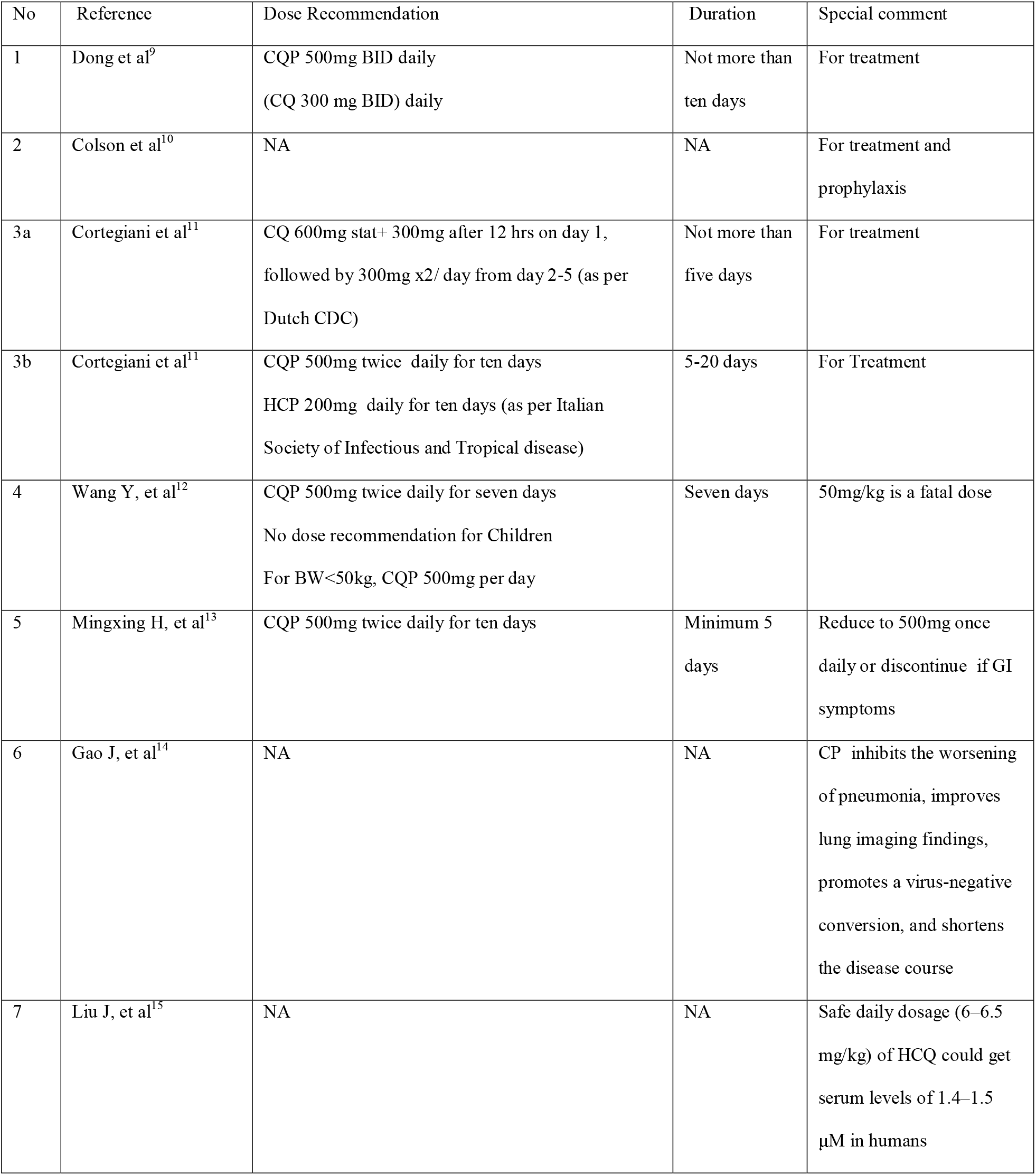
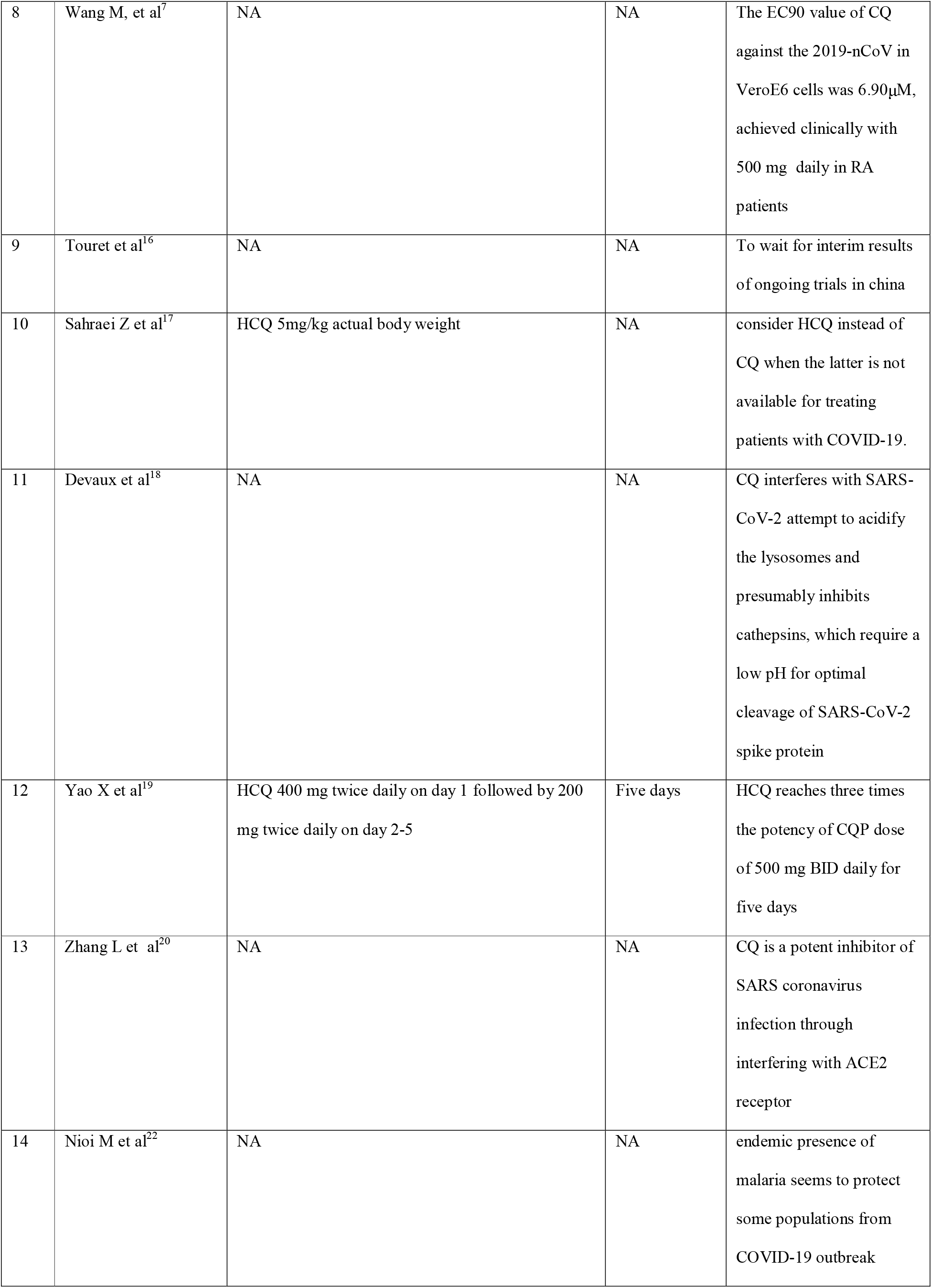
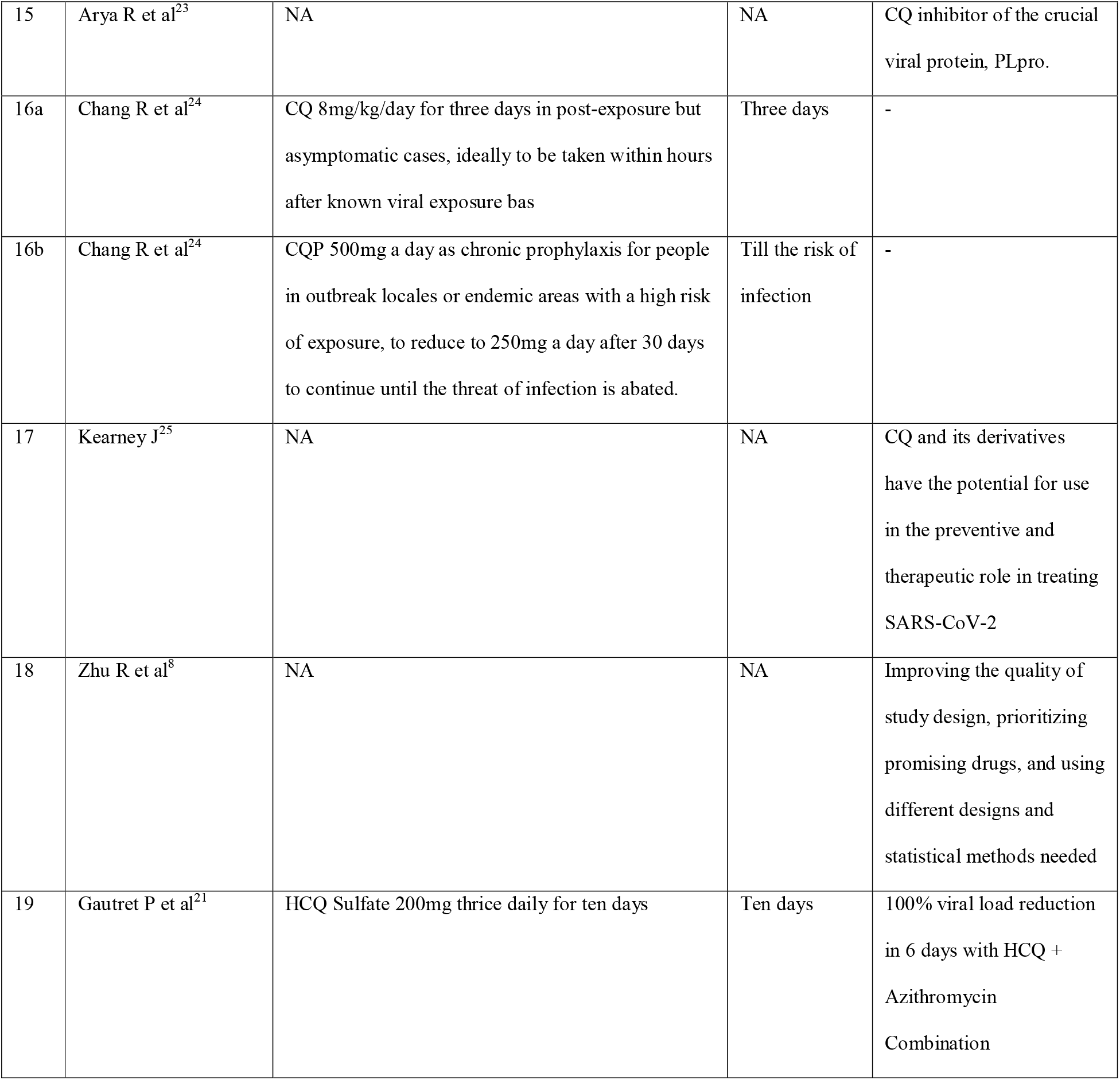
Recommendations in various publications about the role of CQ and HCQ in COVID-19 Infection

In accordance with the “New Coronavirus Pneumonia Diagnosis and Treatment Program (Trial Version 6)” issued by the National Health and Health Commission in China, the recommended dosage of Chloroquine phosphate tablets is 500 mg BID daily for ten days^9^. In case of severe gastrointestinal symptoms, the dose of CQ can be reduced to 500 mg once daily, or can even be discontinued. During the course of CQ treatment, if the throat swab becomes negative and stays negative for three days, the drug withdrawal can be considered. However, the minimum course of CQ treatment needs to be at least five days^13^.

So far, results of more than 100 patients treated with CQ has shown that chloroquine phosphate is superior to the control treatment as it inhibits the worsening of pneumonia, improves the lung imaging findings, promotes a virus-negative conversion, and shortens the course of the disease. Severe adverse reactions to CQ were not seen in these patients. The drug has been recommended for inclusion in the next version of the Guidelines for the Prevention, Diagnosis, and Treatment of Pneumonia Caused by COVID-19, issued by the National Health Commission of the People’s Republic of China.^14^ This would be the first successful use of CQ in humans for treating acute viral disease. However, this information should be carefully considered before drawing any definitive conclusion as no supporting clinical data has been released yet.^16^

As of 23rd February 2020, seven clinical trial registries have been found in the Chinese Clinical Trial Registry for using HCQ to COVID-19 treatment. It has been reported that the safe dosage of HCQ (6– 6.5 mg/kg per day) could reach serum levels of 1.4–1.5 μM in humans. In animals, CQ and HCQ share similar tissue distribution, with concentrations in the liver, spleen, kidney, and lung reaching levels, which are 200–700 times higher than those in the plasma. Therefore a very high concentration in the above tissues is likely to be achieved with the safe dosage of HCQ for inhibiting SARS-CoV-2 infection. This possibility, however, awaits confirmation by clinical trials.^15^ In patients with COVID-19 infection, CQ can interact with lopinavir/ritonavir, Causing QT interval prolongation. Hence HCQ can be considered instead of CQ when the latter is not available for COVID-19 treatment^17^.

HCQ has been found to be more potent than CQ at inhibiting SARS-CoV-2 *in vitro*. HCQ Sulfate 400 mg twice daily for one day, followed by 200 mg twice daily for four more days, has been recommended for treating COVID-19 infection.^19^ Based on CQ’s antiviral mechanisms, its laboratory activity against COVID-19, as well as CQ’s pharmacokinetics based on its use for malaria and autoimmune diseases, safe and potentially efficacious dose regimens have been recommended for protection against COVID-19. This protocol has pre-exposure prophylaxis of 250-500mg CQ daily and post-exposure prophylaxis at 8mg/kg/day CQ for three days.^24^

In a recent trial using HCQ as a single drug and HCQ in combination with azithromycin, the proportion of patients with negative PCR results in nasopharyngeal samples had a significant difference between the two groups. At day six post-treatment, 100% of patients treated with HCQ and azithromycin combination had been virologically cured compared to 57.1% in patients treated with only HCQ and 12.5% in control group.^21^

Contraindications and relative contraindications include NOT to use CQ in people below 18 years and above 65 years, pregnant women, persons allergic to 4-aminoquinoline, patients with hematological disorders or G6PD deficiency, patients with chronic liver or kidney disease, patients with arrhythmia and chronic heart disease, patients with a history of retinal disease or hearing loss, patients known to have a mental illness, Skin diseases like a rash, dermatitis, psoriasis. CQ is known to have drug interactions with various drugs, and expert medical opinion, regarding its interaction with the medicines already taken by patients, is a MUST.

There are some mild side effects of CQ while treating malaria, including headache, dizziness, loss of appetite, etc. The primary adverse reaction of CQ at higher doses is ocular toxicity, which could affect the vision. If the discomfort in the eye or visual abnormality occurs, CQ should be discontinued immediately. Other side effects of CQ include cardiac arrhythmia, drug-induced psychosis, leukopenia, etc. Combining CQ with heparin could increase the risk of bleeding. In patients taking Digitalis, CQ could cause a cardiac block.^13^

For decades, most Europeans visiting malaria-endemic areas have been receiving chloroquine prophylaxis, and they would continue it for two months after their return. The use of CQ for the treatment of malaria has also been in use for long, and HCQ has been used at much higher doses of up to 600 mg/day to treat autoimmune diseases. It is quite difficult to find a product having a better-established safety profile than CQ at such a low cost.^10^ CQ, and its derivatives have shown early promise to treat COVID-19 infection and should be explored as a potential drug in the preventive and therapeutic role to turn the tide in this rapidly evolving pandemic.^25^ Disorderly clinical trials of COVID-19 using traditional Chinese medicine and Western medicine are ongoing but may not be able to give high-quality clinical evidence. In order to death with the health emergency create by COVID-19, the national administration of scientific research should improve management and coordination for improving the study quality based on clinical trial guidelines.^8^

It is also recommended to further research on the role of CQ for COVID-19 prevention. It has also been urged in an article that the above CQ regimens be investigated in parallel with the mass deployment of this drug, without unnecessary regulatory delays, in an attempt to contain the global pandemic as the benefits of CQ far outweigh the risks or costs. ^24^

## Conclusions

The results of in vitro studies and those of recent clinical trials using CQ and HCQ in COVID-19 infection are quite promising. An international strategy for allowing the use of promising drugs like CQ and HCQ for COVID-19 should be made. Considering the low cost, easy availability, and minimal adverse effects, CQ and HCQ should be prescribed to COVID-19 patients under medical supervision. Further trials with this drug will throw more light on its usefulness in COVID-19 infection.

## Limitations

This systematic review had some limitations, including a small sample size of publications and limited data regarding the role of CQ and HCQ in human infection with COVID-19. However, under current emergency circumstances, we believe our study will be beneficial to the scientific community and the general public.

## Data Availability

We have not used an external database. It is a systematic review of the literature.

## Declaration of Competing Interest

N/A

## Funding

No Funding received

## Ethical Approval

Not needed

### Abbreviations

CQ: Chloroquine
CQP: Chloroquine phosphate
HCQ: Hydroxychloroquine
PRISMA: preferred reporting items for systematic reviews and meta-analyses

## Notes

### Competing Interest Statement

The authors have declared no competing interest.

### Clinical Trial

It is a literature review of role of Chloroquine and Hydroxychloroquine in COVID-19 under emergency circumstances. Hence it has not been registered

### Funding Statement

No external funding was received

